# Prediction of the ectasia screening index from raw Casia2 volume data for keratoconus identification by using convolutional neural networks

**DOI:** 10.1101/2024.09.13.24313607

**Authors:** Maziar Mirsalehi, Benjamin Fassbind, Andreas Streich, Achim Langenbucher

## Abstract

**Purpose:** Prediction of Ectasia Screening Index (ESI), an estimator provided by the Casia2 for identifying keratoconus, from raw Optical Coherence Tomography (OCT) data with Convolutional Neural Networks (CNN).

**Methods:** Three CNN architectures (ResNet18, DenseNet121 and EfficientNetB0) were employed to predict the ESI. Mean Absolute Error (MAE) was used as the performance metric for predicting the ESI by the adapted CNN models on the test set. Scans with an ESI value higher than a certain threshold were classified as *Keratoconus*, while the remaining scans were classified as *Not Keratoconus*. The models’ performance was evaluated using metrics such as accuracy, sensitivity, specificity, Positive Predictive Value (PPV) and F1 score on data collected from patients examined at the eye clinic of the Homburg University Hospital. The raw data from the Casia2 device, in 3dv format, was converted into 16 images per examination of one eye. For the training, validation and testing phases, 3689, 1050 and 1078 scans (3dv files) were selected, respectively.

**Results:** In the prediction of the ESI, the MAE values for the adapted ResNet18, DenseNet121 and EfficientNetB0, rounded to two decimal places, were 7.15, 6.64 and 5.86, respectively. In the classification task, the three networks yielded an accuracy of 94.80%, 95.27% and 95.83%, respectively; a sensitivity of 92.07%, 94.64% and 94.17%, respectively; a specificity of 96.61%, 95.69% and 96.92%, respectively; a PPV of 94.72%, 93.55% and 95.28%, respectively; and a F1 score of 93.38%, 94.09% and 94.72%, respectively.

**Conclusions:** Our results show that the prediction of keratokonus based on the ESI values estimated from raw data outperforms previous approaches using processed data. Adapted EfficientNetB0 outperformed both the other adapted models and those in state-of-the-art studies, with the highest accuracy and F1 score.

## Introduction

Keratoconus describes a disorder of the eye characterised by a cone-shaped cornea with thinning and steepening, which typically affects both eyes of a patient with varying degrees of severity and occurs in both males and females [1]. Keratoconus affects about 1 in every 2000 individuals in the general population [2].

There are two main types of corneal imaging: corneal topography and corneal tomography. In corneal topography, the shape of the anterior part of the cornea is shown but in corneal tomography a three-dimensional image of the whole cornea is shown. Optical Coherence Tomography (OCT) is a corneal tomography technique that assesses the delay of reflected infrared light from the anterior segment by comparing it to a reference reflection. This tomography technique is classified into two types: Fourier domain, which uses a stationary mirror and time domain, which adjusts the position of a reference mirror. Another corneal tomography technique is Scheimpflug imaging where a rotating camera is used to produce cross-sectional images [3].

Artificial Intelligence (AI) enables machines to perform tasks associated with human cognition like writing, speaking and seeing. AI can be used in medical specialties dealing with image analysis like ophthalmology. Machine learning is a subset of AI that enables the machine to learn in order to develop its performance. Deep learning, a specialised branch of machine learning, improves the effectiveness of motion recognition, image and speech [4].

In this study, the neural networks were used to predict the ESI of a given scan automatically. This approach is a regression task since the output of the networks is a numerical value. Also, the scans were classified into two classes, *Keratoconus* and *Not Keratoconus*. The *Keratoconus* class represents ectasia and the *Not Keratoconus* class indicates suspicion of ectasia or no ectasia pattern. This approach has an advantage over other approaches where the output is discrete and belongs to a class. With this approach, if two scans are in the *Keratoconus* class, the severity of ectasia can be compared between them by comparing the predicted ESI provided by the model.

In general, data can be utilised as preprocessed data or as raw data. Preprocessed data is altered by software and the details of these modifications may not always be transparent. Moreover, changes in software versions can lead to variations in how data is preprocessed and affect the consistency of results. In contrast, raw data remains unaltered by external software. Therefore, raw data retains its original form across different software versions. This stability in raw data can offer a more consistent and reliable foundation for analysis and model training. To the best of our knowledge, it is the first time that raw OCT data is used for a regression task to predict ESI for the purpose of keratoconus diagnosis. Below we briefly review the current neural network-based approaches to automatically identify keratoconus.

### State of the art

Zhang et al. [5] explored keratoconus diagnosis by employing the CorNet model. The model was trained and evaluated with a dataset of 1786 raw data from the Corvis ST (Oculus, Wetzlar, Germany). Corvis ST is a non-contact device that measures corneal biomechanics by recording dynamic deformation following a rapid air-puff excitation. Keratoconus was diagnosed by using clinical signs such as stromal thinning, Fleischer’s ring and a central K-value greater than 47 dioptres, in addition to other indicators. The CorNet model achieved an accuracy of 92.13%, sensitivity of 92.49%, specificity of 91.54%, PPV of 94.77% and an F1 score of 93.62% on the validation set.

Ruiwei Feng et al. [6] introduced a deep learning method named KerNet for identifying keratoconus and sub-clinical keratoconus using raw data from the Pentacam HR system (Oculus, GmbH, Wetzlar, Germany). This system includes a rotating Scheimpflug camera, which gathers three-dimensional data of the cornea, and a software which is designed to analyse and display the data. The corneal data, exported from the Pentacam HR system, comprised five numerical matrices for each sample. These matrices were considered as five two-dimensional image slices, representing the front and back surface curvatures, the front and back surface elevations and the pachymetry of the eye. 854 samples were used as dataset. KerNet employed a specialised architecture with five branches to handle the matrices individually as input to identify features, which are subsequently combined for prediction. The model achieved an accuracy of 94.74%, with a sensitivity of 93.71%, Positive Predictive Value (PPV) of 94.10% and an F1 score of 93.89%.

Schatterburg et al. [7] introduced a plan for using convolutional neural networks (CNNs) for keratoconus diagnosis based on ESI from data of the SS-1000 Casia OCT Imaging System. The dataset sourced from over 1900 patients and included three-dimensional OCT images of both the anterior and posterior cornea, together with parameters calculated by the Casia software. However, the study did not include evaluation metrics.

Fassbind et al. [8] focused on identifying abnormalities such as keratoconus by employing CorNeXt as a CNN model. In this study, cornea topography maps from Casia2 anterior OCT device were used. The used CorNeXt model is based on the ConvNeXt [9] CNN architecture. To employ ConvNeXt for corneal disease classification, modifications to the architecture were implemented. Measurements of axial refractive power, as well as the elevation of the cornea’s front and back surfaces and its thickness were taken from the scan for every individual cornea and five related maps were created and displayed as grayscale images. ConvNeXt was adapted to include all cornea data by stacking these maps into a five-channel pseudo-image. The dataset included a total of 2182 scans (1552 scans for training, 388 scans for validation and 242 scans for test). The model achieved a sensitivity of 98.46% and a specificity of 91.96% in distinguishing healthy from pathological corneas. For the labeled class of keratoconus, it reached 92.56% accuracy, 84.07% sensitivity, 100% specificity and a 91.34% F1 score.

## Materials and methods

### Convolutional neural network

Artificial neural networks mimic the brain’s processes through nodes and connections. Nodes receive input and send output by processing the input through an activation function, and connections have adjustable weights, which are determined during training and thus allow the network to learn [10]. Activation functions model the way neural electrical signals are passed on to the following neuron [11]. These functions (such as the Sigmoid function, the rectified linear unit and the hyperbolic tangent) add nonlinearity to neural networks [12]. If a network lacks activation functions, its output remains a linear combination of the input regardless of the number of layers; therefore, the intermediate layers become ineffective in contributing to the network’s output. The activation function is located between two layers in a neural network with several layers. A loss function is employed to quantify the difference between the true (observed) values and the values predicted by the model. Intricated optimisers are usually used to with the goal of adapt the model parameters such that the difference between true and predicted outcomes is minimised. Both loss functions and optimisers guide the neural network in learning and adapting to achieve the expected outcomes [11]. However, they are loosely inspired from natural neurons and allow to model functions of arbitrary complexity.

Convolutional Neural Networks (CNNs), as a specific form of neural network, are designed for example for image data [13]. CNNs are designed based on the principles of visual perception, where artificial neurons simulate the behavior of biological neurons and convolution kernels detect different features like receptors [11]. In a convolutional layer, kernels defined by their width, height and a set of weights are applied to the input images to generate feature maps [13].

### Quality criteria

In this study, Mean Square Error (MSE) is used as a loss function for the regression task. MSE is a derivable criterion and having a derivable criterion is essential for gradient descent algorithms, which are used universally to adjust weights in neural networks during training. MSE is defined as Eq (1), where N signifies the number of actual values, which is equivalent to the number of predicted values; *y*_*i*_ represents the actual value at position i and 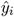 represents the predicted value at the same position [14].

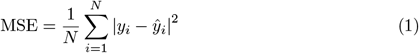

To compare the performance of different prediction models, Mean Absolute Error (MAE) is used, as this measures the average absolute difference between the actual values and the predicted values by the model [11]. Eq (1) illustrates the MAE computation, where N, *y*_*i*_ and 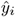 retain the same meanings as in Eq (1) [14].

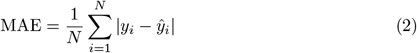

Key metrics for evaluating a binary classifier are derived from the four entries in the confusion matrix. They are crucial for assessing the classifier’s performance. True positive (TP) signifies the count of correctly classified positive samples, such as images with keratoconus correctly identified as having keratoconus. True negative (TN) represents the count of correctly classified negative samples, like images without keratoconus correctly identified as not having keratoconus. False positive (FP) refers to the count of samples that have been incorrectly classified as positive, i.e. in our case, images without keratoconus mistakenly identified as having keratoconus. False negative (FN) indicates the count of samples that have been incorrectly classified as negative, such as images with keratoconus incorrectly identified as not having keratoconus. Table 1 shows the confusion matrix.

**Table 1.** Confusion matrix.

| Actual class | Predicted class |  |
| --- | --- | --- |
|  | Positive | Negative |
| Positive | TP | FN |
| Negative | FP | TN |

In this study, the metrics below are used to assess how effectively the models classify the data into two different categories [15].

Accuracy measures the proportion of correctly classified samples out of the total number of samples in the test dataset. Accuracy is calculated as [15]

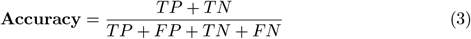

Sensitivity is the proportion of correctly identified positive samples out of all actual positive samples, calculated as [15]

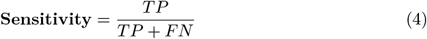

Specificity measures the proportion of correctly classified negative samples out of all samples classified as negative [15]:

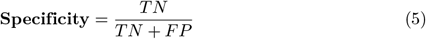

Positive Predictive Value (PPV) is defined as the proportion of correctly classified samples relative to all samples predicted to belong to the positive class [15]:

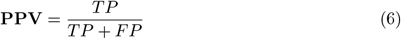

As we are using a threshold on the estimated ESI, a high sensitivity or specificity can be trivially achieved at the cost of a useless low value of the respective other metric. The F1-score finds a balance between these two metrics. The F1 score is defined as [15]

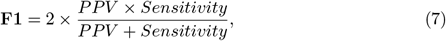

Furthermore, the F1 score has an advantage when dealing with imbalanced datasets, where one class significantly outnumbers the other. In such cases, metrics like accuracy, sensitivity and specificity may not effectively measure how well the model distinguishes between classes. Therefore, the F1 score can be used because it provides a more balanced evaluation of the model’s performance.

The Receiver Operating Characteristic (ROC) curve was analysed to find the best trade-off between sensitivity and specificity for predictions by identifying the optimal threshold, which is the point that maximises the difference between the true positive rate (sensitivity) and the false positive rate (1-specificity). Following this, predicted ESI values were classified into positive and negative classes based on the optimal threshold to compute the confusion matrix values. Predicted ESI values that are equal to or exceed the threshold are considered as *Keratoconus* which indicate the presence of ectasia and those below the threshold are categorised as *Not Keratoconus* which indicate suspicion of ectasia or no ectasia pattern.

### Data

In this study, the data were obtained from patients examined at the eye clinic of the Homburg University Hospital, between February 01, 2021 and September 01, 2023. The data were anonymised at the source and were transferred to us for further processing on October 02, 2023. We were freed from the requirement for ethics approval for the data by the ethics committee of the Saarland medical council (registration number 157/21). Age and sex were not considered important. The Cornea/Anterior Segment OCT Casia2 from Tomey Corporation, made in Japan, was used for data acquisition from patients. This device uses optical coherence tomography with a 1310 nm wavelength laser to measure different parameters like corneal thickness, the depth from anterior surface of the cornea to the anterior surface of the crystalline lens and the depth from the posterior surface of the cornea to the anterior surface of the crystalline lens. The scan range is 13 mm in depth and 16 mm in diameter. Casia2 produces raw data after measurement which has the format of 3dv. The Casia2 device has two modes available: ‘Anterior Segment mode,’ which offers features like the corneal map, and ‘Lens mode,’ which provides lens biometry. In Anterior Segment mode, high-sensitivity measurements of the cornea, angle and intraocular lens can be performed, although it does not allow visualisation of the posterior lens. Conversely, Lens mode provides a simultaneous view of the entire area from the cornea to the posterior lens, however with slightly reduced sensitivity for the cornea. For the detection of keratoconus, the Anterior Segment mode was selected. Each 3dv file related to the corneal map is 36.6 MB in size. For each 3dv file there is an xpf file that contains metadata about the measurement, including the examined eye (left or right), date and time of the examination and the exam protocol name. For each measurement, the ESI is stored in a csv file, which can be exported from the Casia2 software. Ectasia screening identifies keratoconus by independently analysing the shapes of the anterior and posterior cornea. The final diagnosis is based on the results from both assessments. For the anterior cornea, the evaluation focuses on spherical, asymmetry and regular astigmatism components of Fourier analysis. For the posterior cornea, the evaluation focuses on the steepest point of instantaneous power, as well as the asymmetry, regular and higher-order irregular astigmatism components of Fourier analysis. If the analysis area is insufficient for either cornea, the result for that cornea will be marked as ‘N/A’. The final diagnosis is determined by the higher score from either assessment; if both are ‘N/A’, the final result will also be ‘N/A’. If the ESI result ranges from 0 to 4, no ectasia pattern is detected. If the the ESI result is between 5 and 29 suggests a suspicion of ectasia and a result between 30 and 95 indicates clinical ectasia.

We used a Python [16] script to extract 16 images from raw data (3dv file) which originally were stored in a 16-bit unsigned integer format. Each image, with a resolution of 800 pixels in width and 1464 pixels in height, was then saved as a grayscale PNG file. Fig 1 shows a series of 16 resized images, where the height has been reduced to one-third of the original dimension by using a Python script to better represent the realistic shape of the eye. The image preprocessing involved cropping 25% from both the left and right sides to exclude unnecessary eyelid areas and 60% from the bottom to remove regions that did not cover the cornea. After that, the images were resized to a dimension of 224×224 pixels.

**Fig 1.**
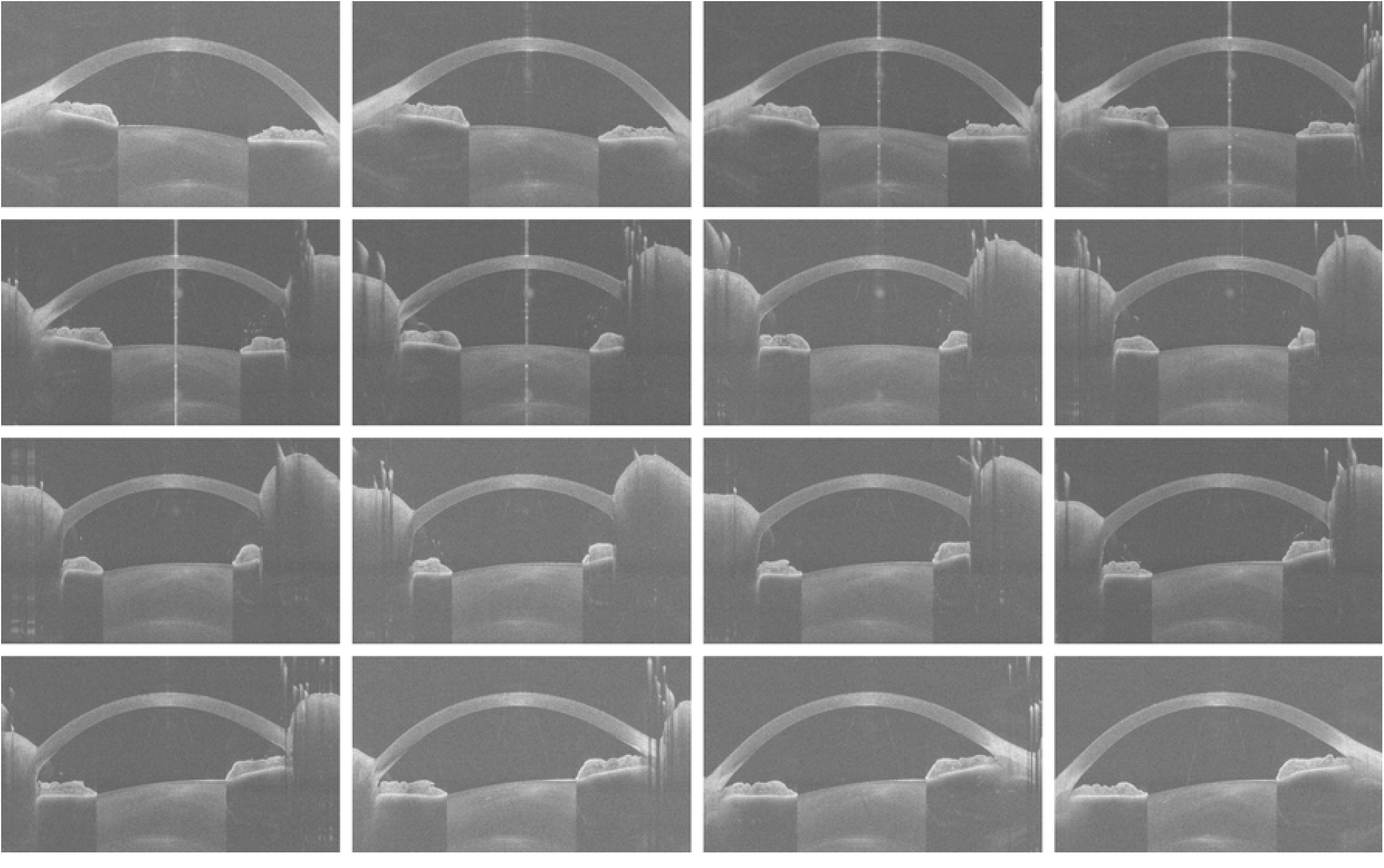
Scaled images of a left eye with an ESI of 0.

### Experimental design and implementation

Since CNNs are suited for detecting objects within images [13], three models (ResNet18, DenseNet121 and EfficientNetB0) were selected based on their performance in the field. ResNet was examined on ImageNet and CIFAR-10 [17], DenseNet was tested on CIFAR-10, CIFAR-100, SVHN and ImageNet [18] and EfficientNet was evaluated on ImageNet and transfer learning datasets, including CIFAR-10, CIFAR-100, Birdsnap, Stanford Cars, Flowers, FGVC Aircraft, Oxford-IIIT Pets and Food-101 [19].

ResNet18 is a variant of the residual network architecture. In residual networks, shortcut connections are used to bypass one or more layers and implement identity mapping which allow their outputs to be summed with the outputs of the intermediate layers [17]. DenseNet121 belongs to the dense convolutional network series. In this type of neural networks, all layers are connected directly with each other which allow them to receive additional inputs from preceding layers and propagate their feature maps to subsequent layers. Unlike residual networks, features are concatenated rather than summed before being forwarded to the subsequent layer [18]. EfficientNetB0 is part of the EfficientNet series. In EfficientNet, the depth, width and resolution of the network are uniformly scaled by a specific set of scaling coefficients [19].

All CNN models were trained from scratch using Python and the PyTorch library [20] on a system equipped with an 11th Gen Intel(R) Core(TM) i7-11700@2.5 GHz processor, 32 GB of RAM and a 64-bit operating system with an x64-based processor. The training proceeded for 100 epochs, during which the validation MSE became stable. The data were divided into disjoint training, validation and test datasets to ensure that the models were trained on one subset, evaluated on another to detect overfitting (where the model fails to apply its learned patterns from training data to unseen data [21]) and finally tested on a separate unseen subset to assess their ability to perform on new data. The batch sizes for the training, validation and test sets were set to 64. From a total of 15457 3dv files, 5817 were selected for training, validation and testing. The files not chosen were excluded due to defects on the cornea, such as keratoplasty. During the training phase, 3689 scans (stored as 3dv file) were used. This represents approximately 63.4% of the total dataset. Similarly, the validation phase involved 1050 scans (accounting for around 18% of the total) and the testing phase consisted of 1078 scans (accounting for 18.5% of the total).

Table 2 presents the distribution of 3dv files which were used for training, validation and testing. The data set is categorised based on ESI, with a threshold of 30, as determined by Casia2. An ESI of 30 or greater indicates the *Keratoconus* class, which signifies clinical ectasia. An ESI below 30 classifies the files as *Not Keratoconus* class, indicating either a suspicion of ectasia or no ectasia pattern detected.

**Table 2.** Data set distribution of 3dv files and classes.

| Data set | Total | *Keratoconus* class | *Not Keratoconus* class |
| --- | --- | --- | --- |
| Train | 3689 | 1486 (40%) | 2203 (60%) |
| Validation | 1050 | 405 (39%) | 645 (61%) |
| Test | 1078 | 429 (40%) | 649 (60%) |

Every set of 16 images from a single 3dv file was stacked together. These stacked images were fed into the models, with the first convolutional layer modified to accept a 16-channel input. The fully connected layer for the output was also modified to produce a single output. Additionally, an extra fully connected layer was included to process the combined features which integrates one feature from the model and two features representing the eye parameters (encoded as a tensor: [1, 0] for the right eye and [0, 1] for the left eye). Each ESI value was used as the label for a set of 16 stacked images in the adapted CNN models. For the training process, the MSE was used as the loss function to minimise prediction errors. Adam is a favoured optimiser for training deep neural networks due to its quicker convergence compared to stochastic gradient descent [22]. Based on [22], AdamW converges faster and generalises better than Adam. In the experiments, the model parameters were optimised using the AdamW optimiser with a learning rate of 0.01 and a weight decay of 0.05. Moreover, a scheduler was implemented to adjust the learning rate on a plateau, with a reduction factor of 0.1 and a patience of 10 epochs.

Fig 2 illustrates the workflow for predicting ESI by using adapted CNN models.

**Fig 2.**
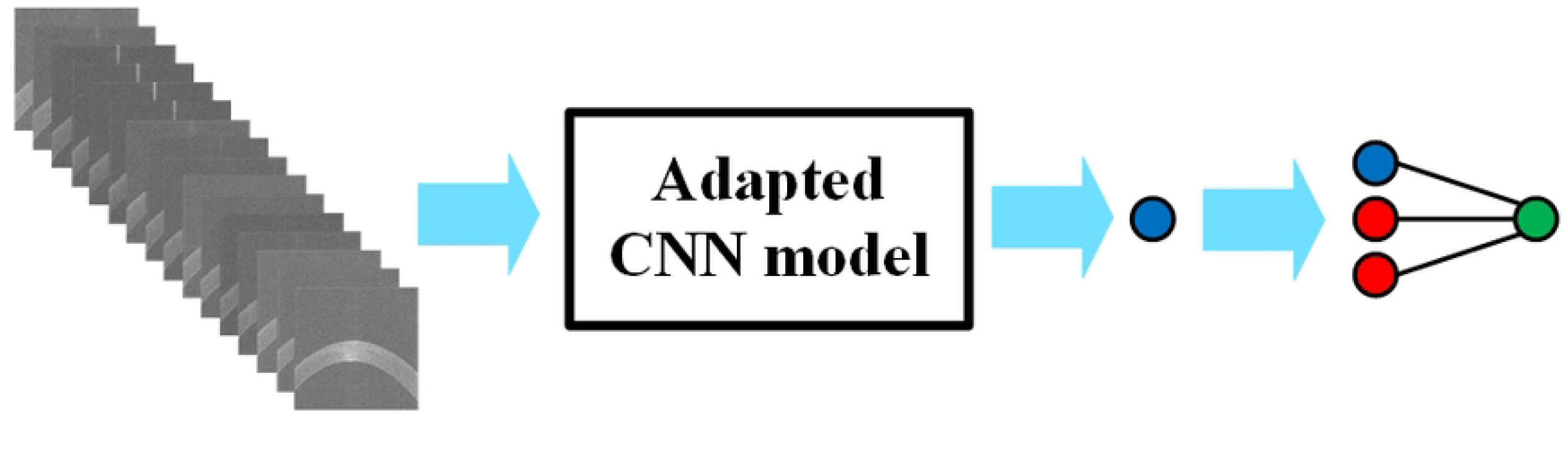
Diagram of the workflow for predicting ESI.

## Results

Table 3 presents the MAE and MSE values, rounded to two decimal places, derived from the evaluation of adapted ResNet18, DenseNet121 and EfficientNetB0 on the test dataset.

**Table 3.**
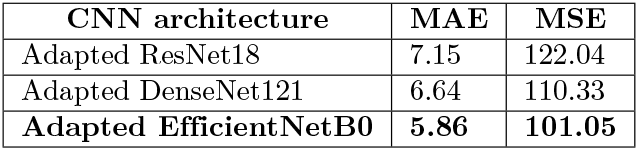
Test set MAE and MSE performance of adapted CNN architectures.

Fig 3 shows kernel density estimates (KDEs) of errors between predicted and actual ESIs for adapted ResNet18, EfficientNetB0 and DenseNet121. These KDE plots represent the distribution of errors, where the error is determined by subtracting the actual ESI from the predicted ESI.

**Fig 3.**
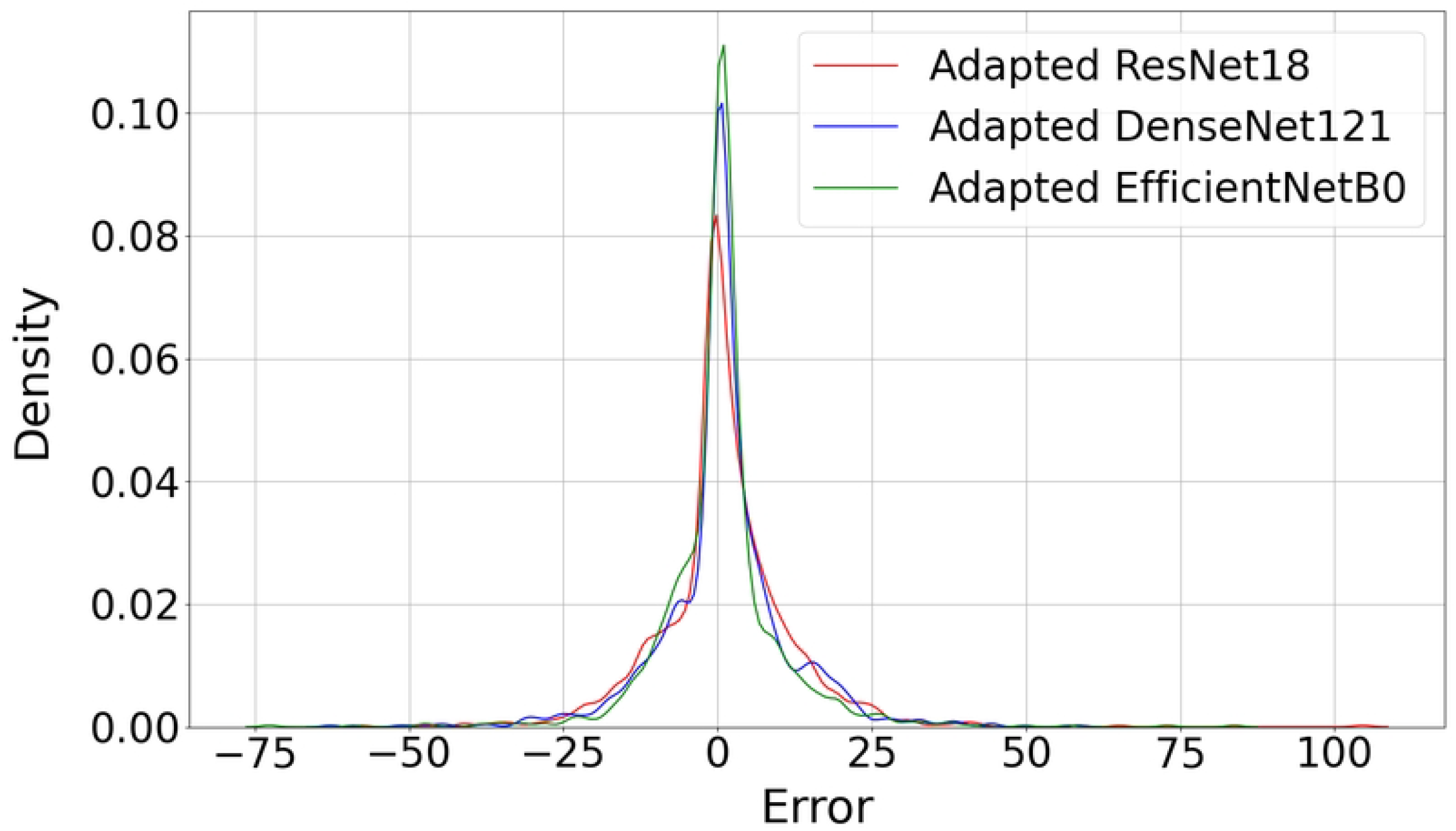
Kernel density estimates of errors between predicted and actual ESIs for different models.

Table 4 provides a summary of the frequency of errors within specified error ranges for the adapted CNN architectures.

**Table 4.** Frequency of errors for CNN architectures within specified ranges.

| CNN architecture | Error range |  |  |  |  |  |  |  |
| --- | --- | --- | --- | --- | --- | --- | --- | --- |
|  | below -10 | -10 to -5 | -5 to -2 | -2 to 0 | 0 to 2 | 2 to 5 | 5 to 10 | above 10 |
| Adapted ResNet18 | 124 | 87 | 74 | 232 | 133 | 134 | 134 | 160 |
| Adapted DenseNet121 | 107 | 100 | 62 | 142 | 267 | 138 | 121 | 141 |
| Adapted EfficientNetB0 | 86 | 113 | 89 | 147 | 298 | 149 | 86 | 110 |

Fig 4 illustrates the correlation between actual ESIs and model predictions for adapted ResNet18, DenseNet121 and EfficientNetB0, respectively.

**Fig 4.**
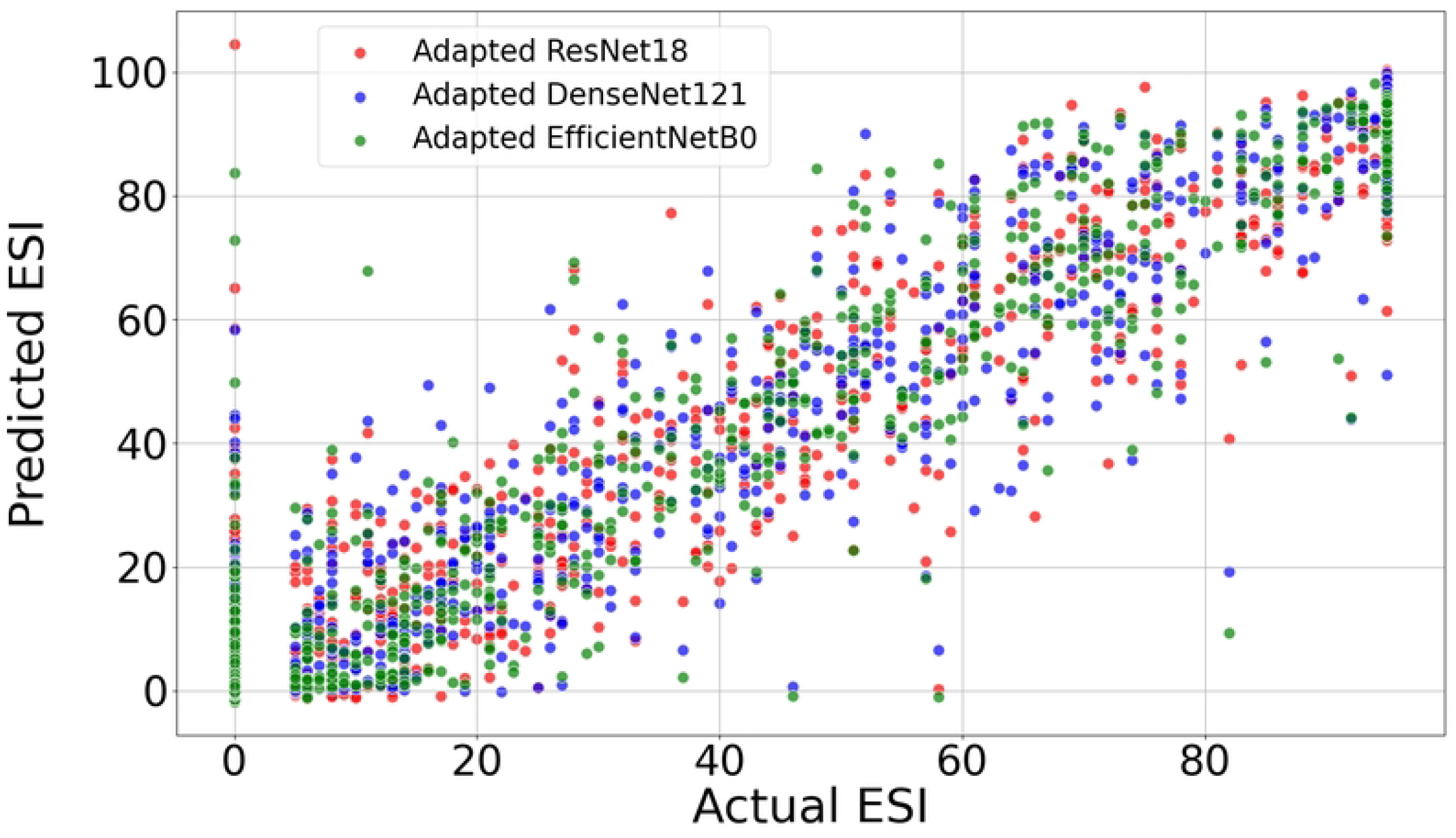
Correlation between actual ESIs and model predictions for different models.

Table 5 summarises the confusion matrices for each of the CNN architectures tested.

**Table 5.** Confusion matrices of CNN architectures.

| CNN architecture | Actual class | Predicted class |  |
| --- | --- | --- | --- |
|  |  | *Keratoconus* | *Not Keratoconus* |
| Adapted ResNet18 | *Keratoconus* | 395 | 34 |
|  | *Not Keratoconus* | 22 | 627 |
| Adapted DenseNet121 | *Keratoconus* | 406 | 23 |
|  | *Not Keratoconus* | 28 | 621 |
| **Adapted EfficientNetB0** | *Keratoconus* | **404** | **25** |
|  | *Not Keratoconus* | **20** | **629** |

Table 6 presents a comparison of classification performance metrics for adapted ResNet18, DenseNet121 and EfficientNetB0 (rounded to four decimal places) with three models of CorNet [5], KerNet [6] and CorNeXt [8] on the test set.

**Table 6.** Evaluation metrics for CNN models.

| CNN Architecture | Metrics |  |  |  |  |
| --- | --- | --- | --- | --- | --- |
|  | Accuracy | Sensitivity | Specificity | PPV | F1 Score |
| Adapted ResNet18 | 0.9480 | 0.9207 | 0.9661 | 0.9472 | 0.9338 |
| Adapted DenseNet121 | 0.9527 | 0.9464 | 0.9569 | 0.9355 | 0.9409 |
| **Adapted EfficientNetB0** | **0.9583** | **0.9417** | **0.9692** | **0.9528** | **0.9472** |
| CorNet [5] | 0.9213 | 0.9249 | 0.9154 | 0.9477 | 0.9362 |
| KerNet [6] | 0.9474 | 0.9371 | None | 0.9410 | 0.9389 |
| CorNeXt [8] | 0.9256 | 0.8407 | 1 | None | 0.9134 |

The optimal thresholds (rounded to two decimal places) for the adapted ResNet18, DenseNet121 and EfficientNetB0 were 26.03, 30.61 and 33.23, respectively.

## Discussion

This study explored the use of three deep neural network architectures (ResNet18, DenseNet121 and EfficientNetB0) for predicting the ESI by using raw data from the Casia2.

Based on the performance metrics presented in the Table 3, the adapted EfficientNetB0 showed the best performance in predicting ESIs on the test dataset. According to Fig 3, the peak around 0 indicates that most predictions from all three models (Adapted ResNet18, DenseNet121, and EfficientNetB0) are very close to the actual ESI values. Also, the plots are centered around zero, which indicates that the errors are symmetrically distributed on either side of the zero error line. Moreover, the adapted EfficientNetB0 model has the highest peak, which indicates that it has the highest proportion of predictions with smaller errors compared to the other two models. Additionally, all models show very low densities of extreme errors (far from zero) which is consistent with Fig 4. According to Table 6, the adapted EfficientNetB0 achieved higher accuracy and F1 score in distinguishing between *Keratoconus* and *Not Keratoconus* classes compared to the two other adapted models and the CorNet, KerNet and CorNeXt models. The higher accuracy and F1 score rates observed for adapted EfficientNetB0 emphasises the potential of this deep learning model in distinguishing between *Keratoconus* and *Not Keratoconus* classes based on the raw data from Casia2.

Future research could explore the applicability of other deep learning architectures beyond the ones evaluated in this study to further enhance performance metrics.

## Conclusion

To the best of our knowledge, this study is the first to use raw OCT data from the Casia2 to predict the ESI. In conclusion, adapted EfficientNetB0 outperformed the adapted ResNet18, adapted DenseNet121 and the models in state-of-the-art studies in distinguishing between *Keratoconus* and *Not Keratoconus* classes. This highlights the effectiveness of this deep learning model in improving diagnostic accuracy and F1 score based on raw data from Casia2 and suggests its significant potential for enhancing ophthalmological evaluations.

## Data Availability

We will upload the minimum dataset to retrieve the relevant results of our paper on a public repository after acceptance of the manuscript.

## Acknowledgments

M M was supported in part by the Rolf M. Schwiete Stiftung under project nr. 2020-024 (https://schwiete-stiftung.com/). The funders had and will not have a role in study design, data collection and analysis, decision to publish, or preparation of the manuscript

